# Circulation of SARS-CoV-2 and co-infection with *Plasmodium falciparum* in Equatorial Guinea

**DOI:** 10.1101/2023.09.12.23295464

**Authors:** Diana López-Farfán, Policarpo Ncogo, Consuelo Oki, Matilde Riloha, Valero Ondo, Pablo Cano-Jiménez, Francisco José Martínez-Martínez, Iñaki Comas, Nerea Irigoyen, Pedro Berzosa, Agustín Benito Llanes, Elena Gómez-Díaz

**Author notes:** Corresponding author: (DL-F).

## Abstract

The impact of COVID-19 in Africa has been a big concern since the beginning of the pandemic. However, low incidence of COVID-19 case severity and mortality has been reported in many African countries, although data are highly heterogeneous and, in some regions, like Sub-Saharan Africa, very scarce. Many of these regions are also the cradle of endemic infectious diseases like malaria. The aim of this study was to determine the prevalence of SARS-CoV-2, the diversity and origin of circulating variants as well as the frequency of co-infections with malaria in Equatorial Guinea. For this purpose, we conducted antigen diagnostic tests for SARS-CoV-2, and microscopy examinations for malaria of 1,556 volunteers at six health centres in Bioko and Bata from June to October 2021. Nasopharyngeal swab samples were also taken for molecular detection of SARS-COV-2 by RT-qPCR and whole genome viral sequencing. We report 3.0% of SARS-CoV-2 and 24.4% of malaria prevalence over the sampling in Equatorial Guinea. SARS-CoV-2 cases were found at a similar frequency in all age groups, whereas the age groups most frequently affected by malaria were children (36.8% [95% CI 30.9-42.7]) and teenagers (34.7% [95% CI 29.5-39.9]). We found six cases of confirmed co-infection of malaria and SARS-CoV-2 distributed among all age groups, representing a 0.4% frequency of co-infection in the whole sampled population. Interestingly, the majority of malaria and SARS-CoV-2 co-infections were mild. We obtained the genome sequences of 43 SARS-CoV-2 isolates, most of which belong to the lineage Delta (AY.43) and that according to our pandemic-scale phylogenies were introduced from Europe in multiple occasions (7 transmission groups and 17 unique introductions). This study is relevant in providing first-time estimates of the actual prevalence of SARS-CoV-2 in this malaria-endemic country, with the identification of circulating variants, their origin, and the occurrence of SARS-CoV-2 and malaria co-infection.

## Introduction

The coronavirus disease 19 (COVID-19) has caused more than 767 million cases and 6,9 million deaths worldwide. However, only a small proportion of these (around 9,5 million cases [1.24%] and 175,339 deaths [2.52%]) have occurred in Africa (data as of July 2023, WHO COVID-19 Dashboard) (1). Moreover, inside Africa, the distribution of cases has been uneven. Most of the reported cases are from a few Northern and Southern countries like South Africa (4.0 million cases), Morocco (1.27 million cases) and Tunisia (1.15 million cases). In comparison, only 634,161 cases account for Central African countries. These data suggest country-specific factors such as reduced transmission or limited testing, raising questions about the real impact of the pandemic in Central African countries.

Genomic SARS-CoV-2 surveillance has been vital in monitoring the emergence, evolution and spread of different variants of concern (VOC). Indeed, several VOC, such as Alpha, Beta, Delta and Omicron, were first detected on the African continent or have spread extensively on it (2–4). Despite the heterogeneous pandemic across Africa, most countries have reported multiple waves of infection (4). Genomic sequencing has also revealed that each wave has been dominated by a different lineage (4,5). Within Africa regional viral population diversity has been observed in the first and second waves and, to a lesser extent, in the third wave. In North and Southern Africa, the first and second waves were dominated by Beta (B1) and Alpha lineages, replaced by Delta and Omicron in the third and fourth waves, respectively. However, different lineages in West, East and Central Africa have dominated the first, second and third waves (4). In addition, other minor lineages have been detected co-circulating and contributing to epidemic waves in all regions of the continent (4,6). In this context, characterising the local diversity of SARS-CoV-2 variants country by country becomes critical.

Equatorial Guinea is located on the west coast of Central Africa and consists of two parts, the mainland and an insular region (i.e., Bioko is the main island). As of April 2023, this country had reported 17,130 cases and 183 deaths, most of them from the insular region (77,2%) (7). Like in other African countries, data about the prevalence of SARS-COV-2 infection in Equatorial Guinea is still scarce. Indeed, to our knowledge, no representative studies exist which evaluate the prevalence of SARS-COV-2 in this country.

After Equatorial Guinea confirmed its first case of COVID-19 community transmission on March 14, 2020, there have been five waves (7). A genomic analysis of SARS-COV-2 sequences revealed that the first wave from April to July 2020 was dominated by the early lineage B.1.192, the second wave from January to April 2021 was dominated by the Beta variant (B.1.351), and the third wave began during July 2021 with the Delta variant (AY.43) (5).

To date, there are deposited 213 SARS-CoV-2 whole genome sequences from Equatorial Guinea on the GISAID database (8). These data correspond, however, to 1.2% of reported cases, which is far from the World Health Organization (WHO) recommendation to sequence at least 5% of COVID-19 cases by country (9). Hence, there is a need to increase the number of SARS-CoV-2 genomes to sequence for a complete assessment of the diversity of variants circulating in the country.

Apart from the direct mortality impact of COVID-19, the pandemic has also resulted in a worsening crisis in controlling several endemic diseases in Africa (10–12). During the first year of the COVID-19 pandemic, malaria cases and deaths in Africa increased from 213 to 228 million cases and from 534,000 to 602,000 deaths (15 million more cases and 68,000 more deaths). This was mainly due to disruption in malaria interventions (13), reaching the worst-case scenario projected at the beginning of the pandemic (14). During 2021, the increase of malaria cases continued but was less marked (2 million more cases) while deaths remained stable (13). This was possible due to the efforts of countries and partners that provided additional funding to deliver essential malaria services (13).

Besides these collateral effects, the circulation of COVID-19 in malaria-endemic countries raises the possibility of co-infection. The frequency and consequences of co-infection at the population level have been assessed in eight African (15,16,25,26,17–24) and one Indian (27) malaria-endemic countries. However, these studies have shown very heterogeneous results depending on the study design and the population or subpopulation sampled (**S1 Table in S1 Text**).

Equatorial Guinea is among the moderate to high malaria transmission countries in the African Region, which accounts for 95% of cases and 96% of malaria deaths globally. To date, no study has examined the occurrence and frequency of SARS-COV-2 and malaria co-infection.

To fill these gaps, in this work, we conducted a cross-sectional study of 1,556 volunteers at six health centres, three in Bioko (island) and three in Bata (mainland), from June to October 2021. All subjects were tested for SARS-CoV-2 by rapid antigen diagnostic tests and for malaria by microscopy. Moreover, swab samples were also taken for SARS-CoV-2 RT-qPCR and whole genome viral sequencing. To our knowledge, this is one of the largest SARS-CoV-2 epidemiological studies conducted in Sub-Saharan Africa. This is also the first to evaluate SARS-COV-2 and malaria co-infection in this endemic malaria country.

## Methods

### Ethics

This protocol was revised by members of the ISCIII/FCSAI at Equatorial Guinea, approved by the Spanish National Research Council (CSIC), the Ethics Committee of Biomedical Research in Andalusia (CCEIBA), and the technical committee of the Ministry of Health and Social Welfare of Equatorial Guinea. The field procedures were carried out by the technical staff of the National Malaria Programme of the Ministry of Health and Social Welfare of Equatorial Guinea. The protocol is in conformity with the declaration of Helsinki and all international regulations about the ethical principles in biomedical research involving human subjects. SARS-CoV-2 and malaria-positive individuals were provided with treatment and assisted by medical professionals. During the screening, local authorities, health centres and volunteers were informed about the objectives of the study and the protocols performed.

### Study design

This study was conducted at six health centres in Equatorial Guinea, three located at Bioko Island (Hospital Regional de Malabo, Centro de Salud Buena Esperanza and Centro de Salud de Campo Yaundé) and three located at the mainland city of Bata (Centro de Salud María Rafols, Centro de Salud La Libertad and Hospital Regional de Bata). 1,556 volunteers were enrolled at the six health centres from ^1st^ June to 4^th^ October 2021 (**S2 Table in S1 text**). Demographic and clinical information collected included age, sex, temperature, symptomatology and pregnancy state. Since COVID-19 vaccination started on March 2021, only non-vaccinated volunteers were eligible for the study.

Prior to taking any samples, participants were required to sign the informed consent form created for the study. For each participating subject, one drop of blood was obtained by capillary puncture of the fingertip. This blood was used for malaria detection by microscopy. When there was consent, a nasopharyngeal sample was also collected for rapid antigen detection of SARS-CoV-2. For SARS-CoV-2 positive antigen test volunteers, a second nasopharyngeal sample was taken, and the swab was stored in pre-filled tubes containing 0.5 mL of inactivating lysis buffer (AVL, QIAGEN) for molecular testing of SARS-CoV-2 and whole genome viral sequencing. These samples were kept at 4°C for 24h and at −20°C or −80°C for extended storage and subsequent analysis.

### Malaria and SARS-CoV-2 diagnosis

Malaria was detected by microscopy following standard methods (28). Blood thick smears were screened for *Plasmodium falciparum* asexual and gametocyte stages in the blood.

Antigen detection of SARS-CoV-2 was performed from individual nasal swabs samples using the Panbio™ COVID-19 Ag Rapid Test (Abbot) following the manufacturer’s instructions. For SARS-COV-2 molecular detection, RNA was extracted from 140 µL of the AVL preservative solution using the QIAamp viral RNA extraction kit (QIAGEN) following the manufacturer’s instructions. RT-qPCR analysis was performed using the one-step Direct SARS-CoV-2 Realtime PCR Kit (Vircell S.L) following the manufactureŕs instructions, reactions were performed in 20 µL final volume (5 µL of RNA sample and 15 µL of RT-PCR mix) using a CFX96 Real-Time PCR Detection System (Bio-Rad, Hercules, CA, USA). Cycle threshold (Ct) values were analysed using the BIORAD CFX manager software. Gene targets identified with this kit were the *nucleocapsid* (N), the *envelope* (E) and the human *RNase P*. Samples were considered positive when the N and E target genes had a cycle threshold (Ct) < 40. Positive and negative controls were always included.

### Statistical analysis

Association between variables was tested with the Wilcoxon Sum Rank tests (for continuous quantitative variables) and the Kruskal-Wallis chi-squared test.

### Whole genome sequencing and phylogenetic analysis

RNA from SARS-CoV-2 positive samples with a Ct value ≤ 35 was sent for sequencing to the Instituto de Biomedicina de Valencia (IBV-CSIC). RNA was retrotranscribed into cDNA, and SARS-CoV-2 complete genome amplification was performed in two multiplex PCR, accordingly to openly available protocol developed by the ARTIC network5 using the V4.1 multiplex primers scheme (artic-network n.d.). Two resulting amplicon pools were combined and used for library preparation. Genomic libraries were constructed with the Nextera DNA Flex Sample Preparation kit (Illumina Inc., San Diego, CA) according to the manufacturer’s protocol with five cycles for indexing PCR. Whole genome sequencing was performed in a MiSeq platform (2 × 150 cycles paired-end run; Illumina). Sequences obtained went through an open-source bioinformatic pipeline based on IVAR6 (29). The different steps of the pipeline are as follows: 1) removal of human reads with Kraken (30); 2) filtering of the fastq files using fastp v 0.20.1 (31) (arguments: –cut tail, –cut-window-size, –cut-mean-quality, -max_len1, -max_len2); 3) mapping and variant calling using IVAR v 1.2; and 4) quality control assessment with MultiQC (32). Consensus sequences generated by this pipeline were aligned against the SARS-CoV-2 reference sequence (33) with MAFFT (34). Problematic positions were masked using the mask_alignment.py script from the repository maintained by Rob Lanfear7. Lineages and clade nomenclature were assigned using Pangolin (35) and Nexclade online tool (36). The phylogenetic tree was generated with Nextclade v2.5.08, downloaded in JSON format and visualised using Auspice v2.37.3 online tool9 (37,38).

### Genome-based epidemiological analysis

A genome-based epidemiological analysis was performed on the sequenced Guinea Equatorial samples following the pipeline described in Martínez-Martínez et al (39). We used a pandemic-scale phylogenomics analysis tool called UShER (40) to place the newly-sequenced samples in a prebuilt phylogeny. For this purpose, we downloaded an existing SARS-CoV-2 phylogeny (eTree) composed of 15.649.343 sequences uploaded in GISAID (8) on July 27th, 2023 from the UCSC ftp server (ftp:apache/cgi-bin/hgPhyloPlaceData/wuhcor1). This method avoids subsampling the dataset by the country of origin or the sampling date, maintaining the broadest possible context.

Using Robert Lanfear’s global_profile_alignment.sh (41), we aligned the Guinea Equatorial sequences against the Wuhan reference sequence (MN908947.3). We used the faToVcf tool from the UShER toolbox to obtain the VCF file, and used this file as input along with the eTree to place the new 43 sequences.

To speed up the analysis, we extracted the Beta and Delta variants’ clades from the eTree using matUtils (42) after identifying the parental nodes using the Taxonium viewer (43). Finally, using a custom code developed in R, we identified the likely country/region of origin and the period of the introductions. To do so, we first defined 3 possible categories for our sequences: transmission group, if they fell in a node in which >60% samples were from Equatorial Guinea; unique sequence, if they fell in a node with lower presence of the country and the length of their branches is greater than 0; and unclassified, if they had no branch length. Then, we searched for the closest sequence with the minimal genetic distance in SNPs that was collected within a period between the 3 months prior to the sampling date, and the sampling date minus 10 days. All the commands used in this analysis are available at the GitLab repository along with the custom code (https://gitlab.com/tbgenomicsunit/covid-guinea).

### Ethics Statement

The studies involving human participants were reviewed and approved by Ethics Committee of the Spanish National Research Council (CSIC), the Ethics Committee of Biomedical Research in Andalusia (CCEIBA), and the technical committee of the Ministry of Health and Social Welfare of Equatorial Guinea. Participants were required to sign the informed consent form created for the study. As the study involved children, a signed declaration of conformity for participating was obtained from their parents and/or legal representatives.

## Results and Discussion

Compared to the rest of the world, the impact of COVID-19 in Africa has been relatively low in terms of severe disease incidence and mortality rate. Several specific socio-ecological and economic factors (i.e., warm weather, younger population, lower population density and mobility, limited testing capacity and previously trained immunity by past infectious diseases) have been proposed to explain the apparent lower transmission (44,45). Other reports, however, suggest that this impact has been vastly underestimated, especially in low-income countries, due to insufficient testing and uneven genomic surveillance. This is probably the case in Equatorial Guinea, where no comprehensive studies assess the prevalence of SARS-CoV-2 and the number of viral genomes available is still very low.

Apart from the noticeable direct effects, the COVID-19 pandemic severely affected the global fight against several endemic infectious diseases, namely VIH, tuberculosis and malaria, mainly because of an interruption and reduction in their control programmes (10–12). But recent data points towards a more direct biological interaction between these diseases In the case of malaria, it has been suggested that severity of COVID-19 infection in malaria-endemic regions may be influenced by co-infection or previous malaria exposure (20,27). Some studies also suggested a possible cross-immunity due to common immunodominant epitopes between the malaria parasite and SARS-CoV-2 (46–48). Prevalence studies of SARS-CoV-2 and co-infection with malaria in sub-Saharan countries are therefore essential to understand the real impact of the pandemic in the African continent.

This is the first comprehensive study of SARS-CoV-2 prevalence in Equatorial Guinea and the first to evaluate malaria co-infection. For this purpose, 1,556 volunteers were recruited at six Equatorial Guinea health centres for four months in 2021 (from June to October) (**S2 Table in S1 Text**). The study was conducted in three health centres on Bioko Island (the site of the country capital, Malabo) and three health centres in the mainland city of Bata. Of the participants, 72.6% were female and 27.4% were male. Samples were distributed to four age groups as follows: A. 5-12 years old (16.8%), B. 13-20 years old (20.8%), C. 21-40 years old (46.2%) and D. >40 years (16.3%). The distribution of positive cases by age and gender is detailed in **Table 1**.

**Table 1.**
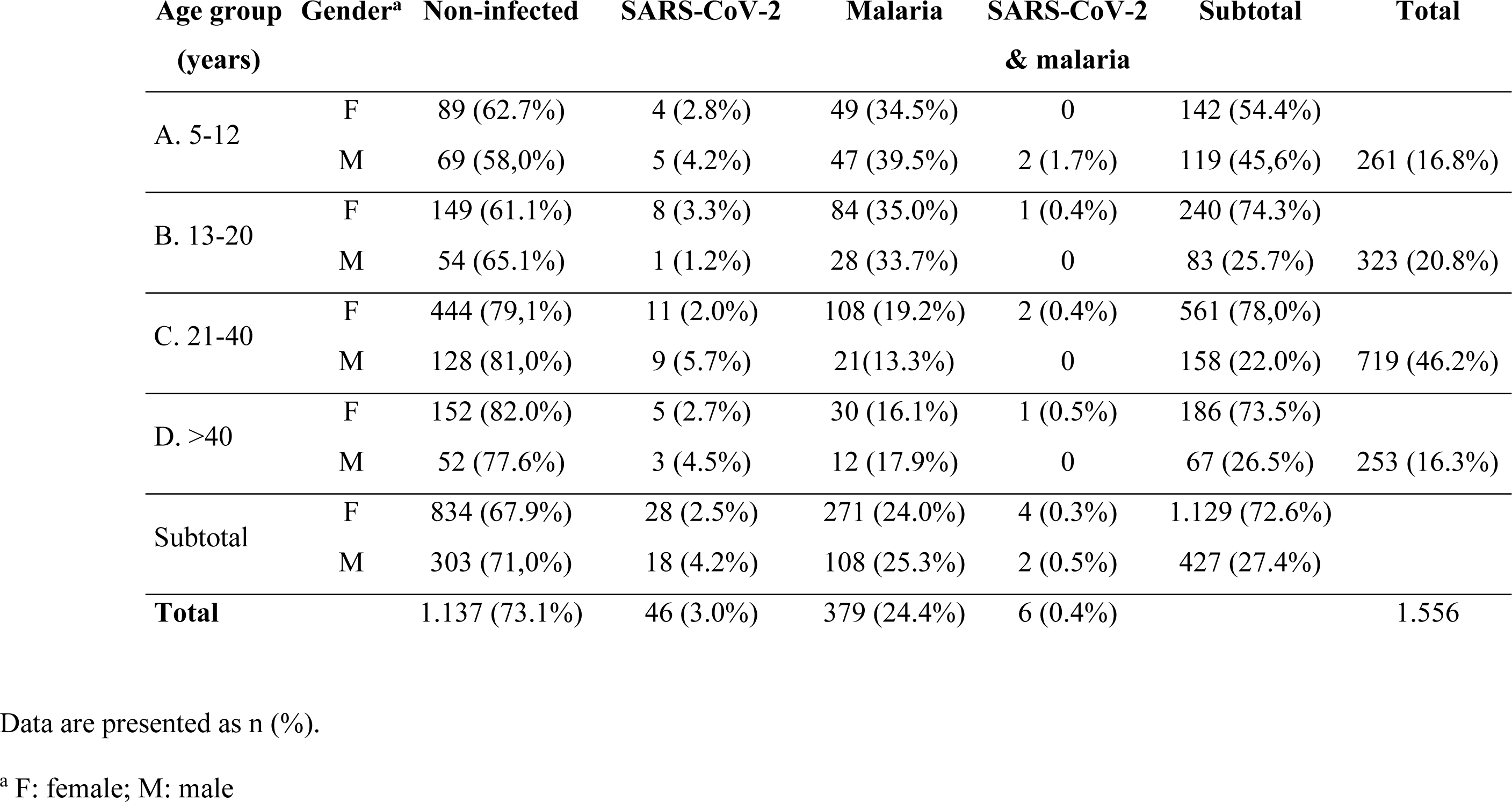
Demographic characteristics and distribution of positive cases in the whole sampled population by age and gender.

For this sampling period and subjects, a total of 46 individuals tested positive by SARS-CoV-2 antigen rapid diagnostic tests, resulting in an overall prevalence of 3.0% (95% CI 2.1-3.8). However, most of the positives were in the health centres of Bioko Island, resulting in a local prevalence of 6.7% (43 positives out of 643 samples) (**S2 Table in S1 Text**). This agrees with the official data from Equatorial Guinea authorities reporting that most SARS-CoV-2 cases were from Bioko Island (7), the site of the capital. SARS-CoV-2 positive cases were found at similar prevalence in all age groups (**Fig 1A**) and there were no significant differences between males and females (W = 39812, *p*-value = 0.07) (**Fig 1B**). However, a temporal trend for SARS-CoV-2 positives increased progressively over the sampling period to reach a peak of 10% of prevalence in September 2021 (**Fig 2**). This trend agrees with the official reports for this country that recorded 4,739 COVID-19 cases by September 2021 (49), coinciding with our sampling period with a peak of cases. Regarding the symptomatology of these cases, most of the SARS-CoV-2 positive cases were mild symptomatic (91.3%), with fever as the most common symptom (**S3 and S4 Tables in S1 Text**).

**Fig 1.**
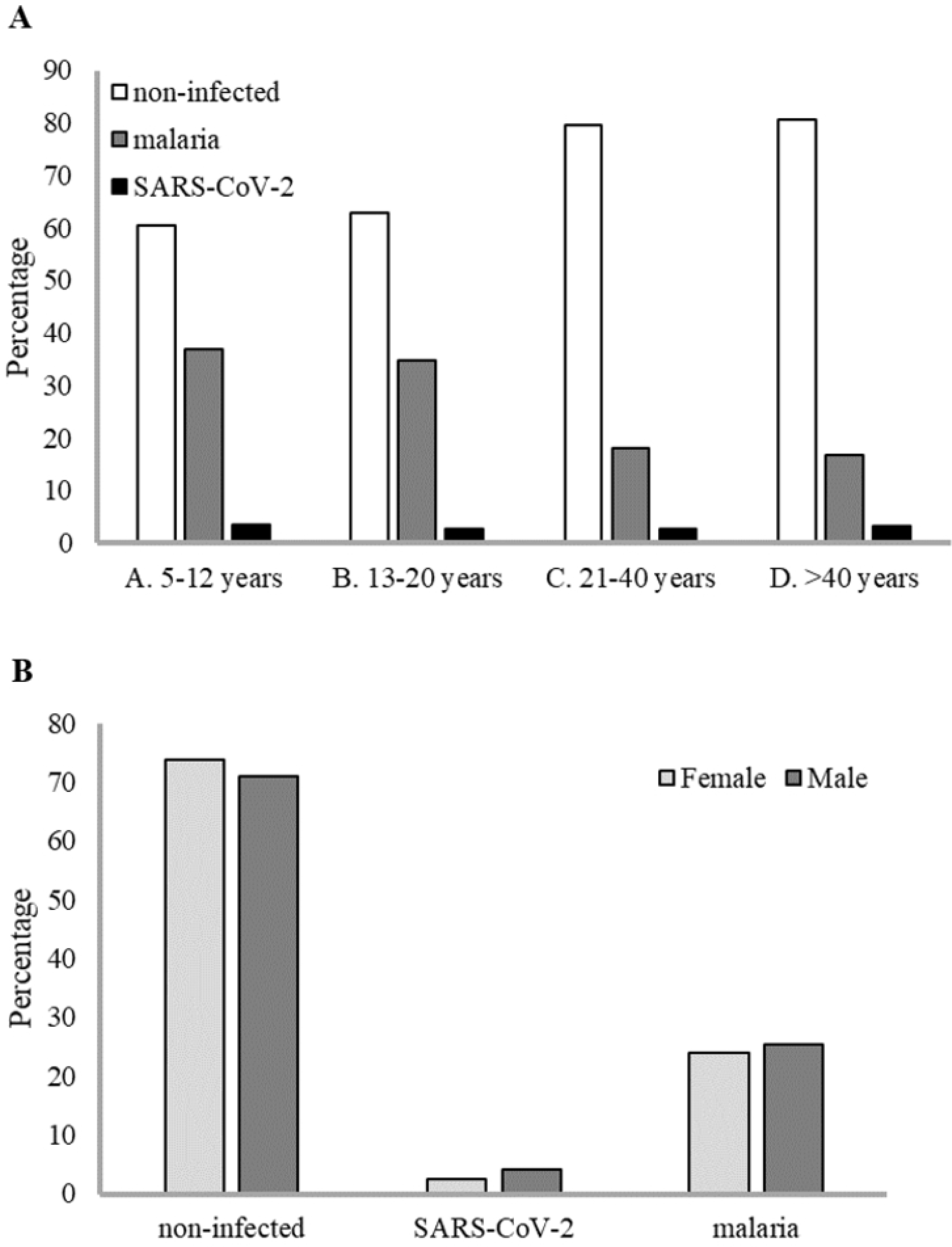
Prevalence of SARS-CoV-2 and malaria cases by age group and gender. (A) Prevalence by age group. (B) Prevalence by gender.

**Fig 2.**
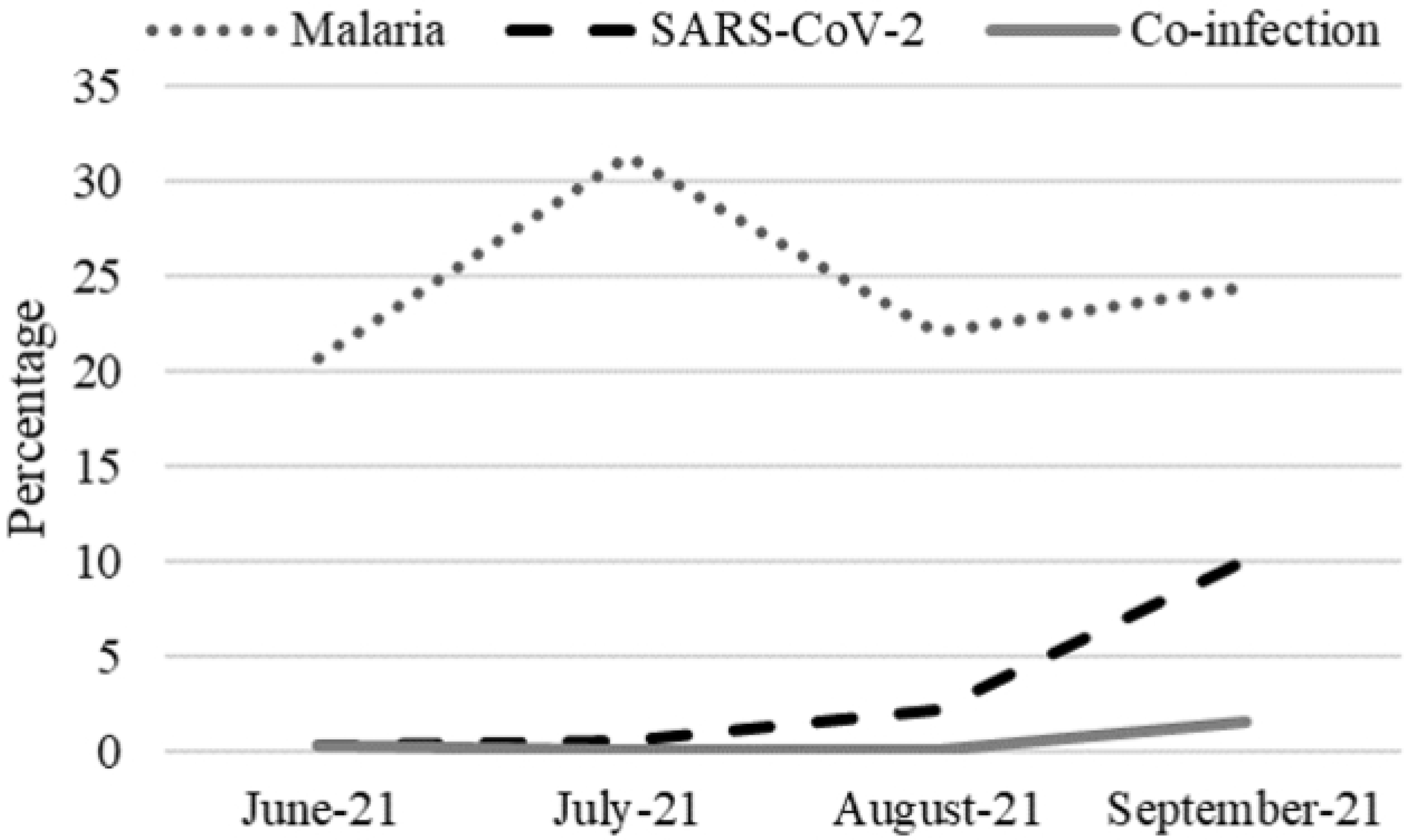
Prevalence of SARS-CoV-2 and malaria over the study period.

Nasopharyngeal swabs available of 43 SARS-CoV-2 positive samples were confirmed positive by RT-qPCR with high viral loads, and cycle threshold values were in the range of 7-29. The RNA of these 43 PCR-positive samples was sent for whole genome sequencing. After genome assembly and quality control, good quality genomes were obtained from 40 samples and medium/low quality genomes from three samples. All the sequences were assigned to lineages using the dynamic lineage classification method, Phylogenetic Assignment of Named Global Outbreak Lineages (PANGOLIN) (35). Phylogenetic analysis revealed that most of the sequences belonged to the lineage AY.43 (38 samples), however, other minor lineages were found, like AY.36 (3 samples) and B.1.351 (2 samples) (**Fig 3 and S5 Table in S1 text**). Our results are in line with a recent genomic study that described the SARS-CoV-2 lineages for the first three waves of the pandemic in Equatorial Guinea from February 2020 to October 2021 (5). That study reported that the first wave from April to July 2020 was dominated by the early lineage B.1.192, the second wave from January to April 2021 was caused by the Beta VOC (B.1.351), and the third wave that began in July 2021 was dominated by the Delta VOC (AY.43).

**Fig 3.**
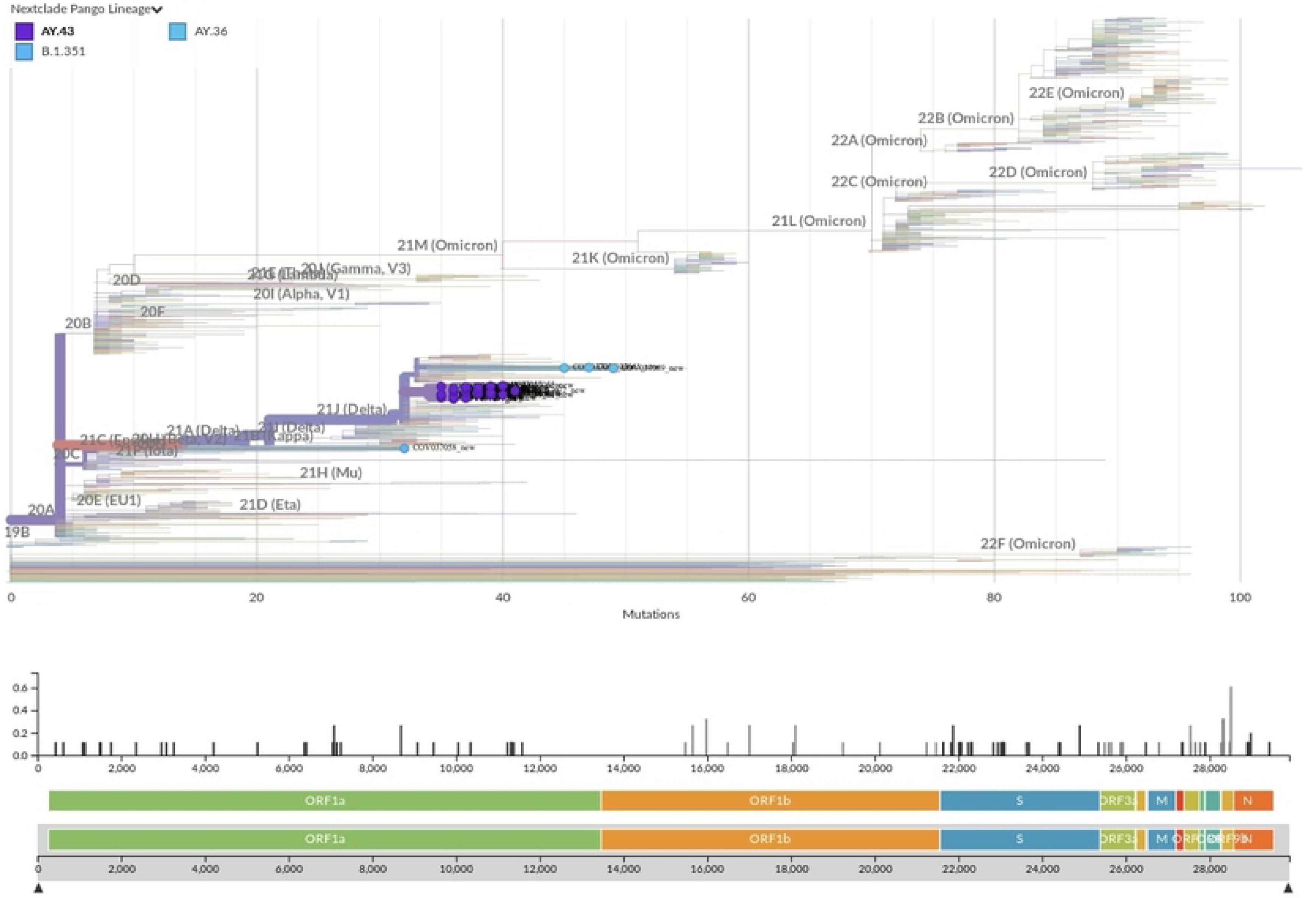
Phylogenetic tree with the 43 new SARS-CoV-2 genomes from Equatorial Guinea. Equatorial Guinea genomes placed on a reference tree of 1,495 published genomes from all over the world. Phylogenetic tree generated with Nextclade online software v2.5.0 (https://clades.nextstrain.org) (accessed on 01 Jun 2023) and visualized using Auspice v2.37.3 (https://auspice.us/).

Our sampling period began in June 2021, when a few positive cases were detected and coincided with the end of the second wave (**Fig 2**). From the end of June to the first half of July, the two genomes available were identified as Beta VOC (B.1.351). The lineage B.1.351 was first identified in South Africa in September 2020 and subsequently defined as a variant of concern (VOC) (3). Since then, it spread to other African countries (50). The earliest sequence of this lineage reported from Equatorial Guinea was in January 2021 (51).

However, the peak of our sampling mostly overlaps with the third wave of the pandemic, between August and September 2021 (**Fig 2**). For that period, most isolated sequences were identified as the Delta variant (AY.43), the VOC that dominated the third wave in Equatorial Guinea. Delta variant was first detected in India in late 2020 (52), from where it was spread to more than 170 countries, including 37 African territories (4). It has been reported that most of the introductions of this variant into Africa (72%) can be attributed to India (4).

The other minor lineage found in this study was AY.36, corresponding to samples collected at the end of August. This is an uncommon Delta lineage that was dominant in Nigeria by June-August 2021 (80% cases) but that represented less 0.5% of Delta cases globally as of September 2021 (6). Its introduction in Nigeria was inferred due to a single introduction during April 2021 and the subsequent spread in the country (6).

To better determine the likely origin and introduction period of the Equatorial Guinea sequenced samples, we performed a genome-based epidemiological analysis using pandemic-scale phylogenies (53). The results are summarized in **Fig 4 and Table S6**, our script identifies 17 unique introductions and 7 transmission groups or clusters, whereas 11 out of 43 samples remain unclassified. Regarding the origin of the introductions, both the transmission groups and unique introductions are mostly of European origin (71.4%, 5 out of 7, and 47.1%, 8 out of 17; respectively), in these cases we cannot identify the exact country since many potential origins were found.

**Fig 4.**
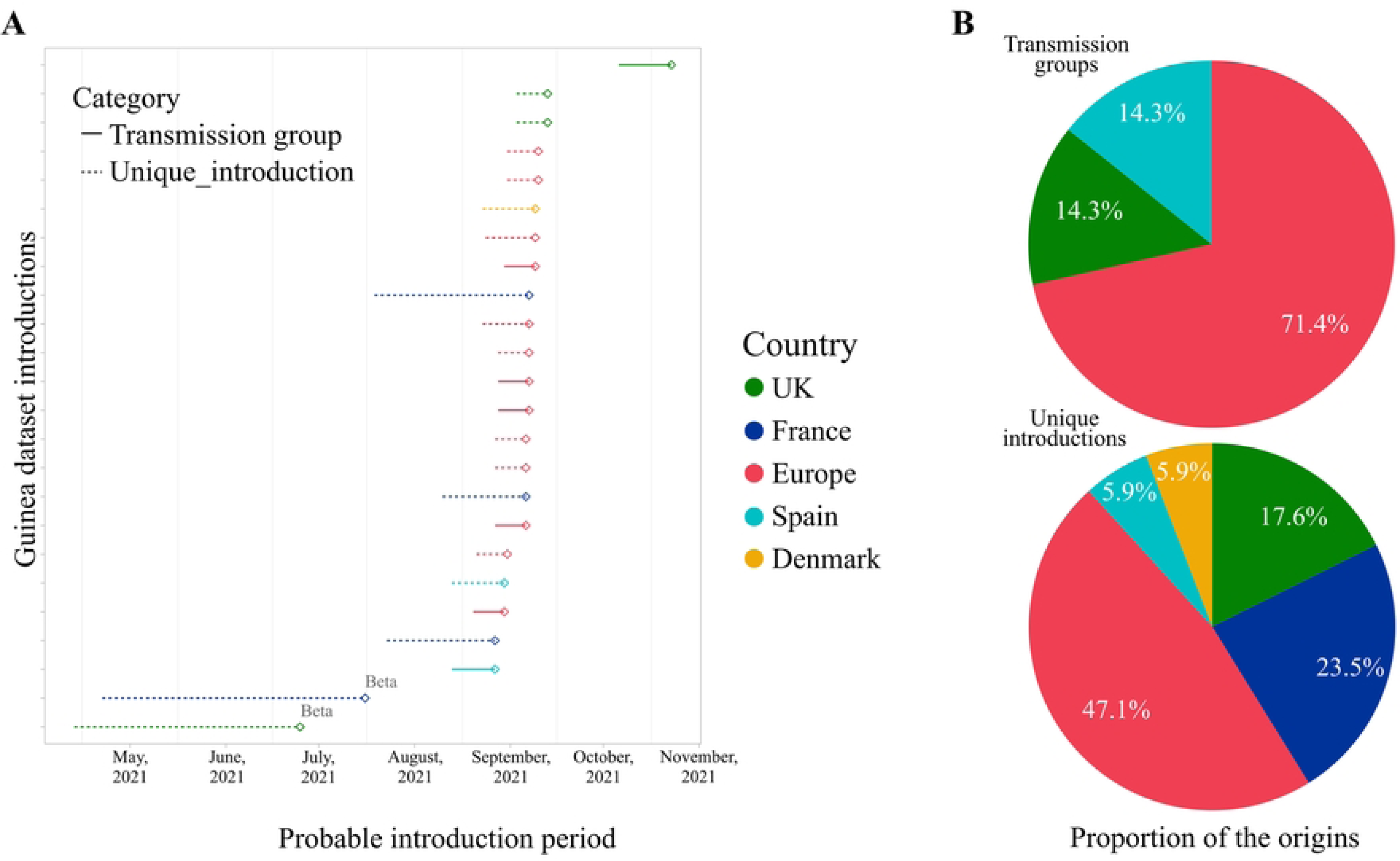
Origin and introduction period of the Equatorial Guinea sequenced samples. (A) Dot plots display introduction periods for transmission groups and unique sequences. Each dot and its segment represent an introduction and its estimated time interval of entrance in the country, coloured by the origin. (B) Pie charts represent the proportion of introductions for transmission groups (top pie) and unique introductions (bottom pie).

In those cases, where the country of origin was identified, we found one transmission group from the United Kingdom (UK) and another from Spain. Concerning 9 unique introductions identified by country, they come from France (4/17, 23.5%), UK (3/17, 17.6%), Spain (1/17, 5.9%) and Denmark (1/17, 5.9%). The introductions from Spain correspond to the three AY.36 samples, the two Beta (B.1.351) samples came from UK and France, while the 27 identified AY.43 samples belong to Europe (16/27), France (7/27), UK (3/27) and Demark (1/27). Collectively, these results point to that the introduction of SARS-CoV-2 to Equatorial Guinea was mainly from outside Africa, not from neighbouring countries.

Regarding the estimated time interval of the introductions, both Beta introductions present a wider interval when compared with Delta, with a range of 2 to 3 months of uncertainty (**Fig 4**). However, this may be due to the large-scale sequencing effort of Delta compared to Beta (∼39 K Beta sequences vs. ∼ 5 M Delta sequences in GISAID). In our analysis, most Delta introductions occurred between mid-July and mid-September, in concordance with the Delta wave seen at a global scale.

In relation to malaria incidence in Equatorial Guinea, 379 individuals were positive by microscopy out of 1,556 tested (24.4% [95% CI 22.2-26.5]). No significant sex difference was observed (W = 226149, *p*-value = 0.6) (**Fig 1B**). However, there were significant differences between age groups in malaria prevalence (Kruskal-Wallis chi-squared = 64.812, df = 3, *p*-value < 0.001), with malaria mainly affecting children aged below 12 years (36.8% [95% CI 30.9-42.7]) and teenagers between 13-20 years old (34.7% [95% CI 29.5-39.9]) (**Fig 1A)**. The majority of the malaria-positive individuals were symptomatic (92,3%), and the most common symptom was fever (≥37.5), but also included other mild to moderate symptoms such as headache, weakness, muscle and joint pain, cough, dizziness, vomiting, diarrhoea and anorexia (**S3 and S4 Tables in S1 text**). No comorbidities were reported among the malaria-positive cases.

When considering these two diseases together, we report six cases of confirmed SARS-CoV-2 co-infection with malaria. Of them, two were children (8 and 10 years old), one was a teenager (14 years old), and three were adults (25, 33 and 41 years old) (**Table 1**). Most of the co-infections were detected during the peak of SARS-CoV-2 cases in September 2021 (**Fig 2**). The sequenced co-infection cases were the dominant lineage AY.43 (4 samples) and B.1.351 (1 sample) (**S5 Table in S1 Text**). One case was not sequenced because the swab sample could not be obtained. Five co-infection cases were symptomatic, the symptoms registered for three cases were mild and included fever, weakness, headache and dizziness. For the other two cases the symptoms were not specified (**S3 and S4 Tables in S1 Text**).

The overall co-infection prevalence reported in this study is low, 0.4% (6/1,556), however, if we calculate the co-infection prevalence among the malaria-positive subpopulation, it corresponds to the 1.6% (6/379), and the co-infection prevalence among the COVID-19 subpopulation is 13% (6/46). One possible explanation for the low rate of co-infections in our study could be the small sample size for the age group of adults >40 years (16.3%, 253/1,556) and children of 5-12 years (16.8%, 261/1,556), the groups most affected by COVID-19 and malaria, respectively. Indeed, in our previous study carried out in Burkina Faso, most of the co-infections were detected in children and teenagers (15), while in another study of Uganda, the highest prevalence of co-infection was in the age groups of 0-20 years and above 60 years (20).

Apart from this study, SARS-CoV-2 and malaria co-infection population studies have been conducted on nine malaria-endemic countries with heterogeneous results depending on the study design and the population or subpopulation studied (**S1 Table in S1 Text**). Most studies have evaluated malaria co-infection prevalence among COVID-19 confirmed patients reporting a wide range of prevalence from 0.63% to 100% (17–21,23,26). Among the few studies that have analysed a random population sample, the low co-infection prevalence that we found in Equatorial Guinea (0.4%) is in the range with what has been reported from Nigeria (0.32%) (16), India (0.73%) (27), Burkina Faso (1.9%) (15), Angola (1.9%) (25) and Gabon (2.2%) (24).

Finally, the low co-infection rate in this and other studies is supported by our previous work in which we explored “*in vitro*” co-infection of human erythrocytes by *Plasmodium* and SARS-CoV-2. In that study, we reported a low entry of SARS-CoV-2 and a negligible interaction with *Plasmodium falciparum*, so the presence of either pathogen does not affect SARS-CoV-2 or *Plasmodium* infection (54). Our findings in mature erythrocytes were in contrast to other studies that reported direct SARS-CoV-2 infection of human erythroid progenitors causing impaired haemoglobin homeostasis, anaemia and other coagulopathies observed in COVID-19 patients (55,56).

The low SARS-CoV-2 prevalence reported in Equatorial Guinea and the low case severity would support the hypothesis of protection by cross-immunity or trained immunity due to co-occurring endemic diseases such as malaria (46–48). In this study, however, we cannot conclude the consequences of malaria and COVID-19 co-infection since the clinical course for co-infection cases was not systematically recorded. Some other studies have addressed the clinical consequences of malaria and COVID-19 co-infection but with contradictory results. Recent retrospective studies of COVID-19 patients have found that malaria co-infection is associated with prolonged hospitalisation (26) and had greater mortality risk compared with just SARS-CoV-2 infection (23). However, other studies report that patients with SARS-CoV-2 and malaria did not seem to have a worst disease outcome, but previous malaria exposure seems to be related to less frequency of severe COVID-19 (20). In another study, COVID-19 patients co-infected with malaria had significantly faster recovery compared to those not co-infected (27). All this considered, additional retrospective and prospective studies are required to better understand the clinical implications of SARS-CoV-2 and malaria co-infections.

## Conclusion

The present study expands on the limited knowledge about COVID-19 prevalence in Central Africa. Our results provide further insights into SARS-COV-2 variant circulation, transmission and evolution in Africa, and confirmed that by June to October 2021, the Delta variant AY.43 was dominating the SARS-CoV-2 infections in Equatorial Guinea, together with a few minor variants. This study also challenges the assumption of Africa as a source of new variants that subsequently spread globally. Finally, we report SARS-CoV-2 and malaria co-infection cases but at low frequency in the population. Importantly, co-infection cases did not worsen symptoms or severity.

## Data Availability

The datasets generated for this study can be found in the European Nucleotide Archive (ENA) database (https://www.ebi.ac.uk/ena/browser/home) with project ID PRJEB65711 and sample IDs ERS16307344-ERS16307386.

## Acknowledgments

We would like to thank all volunteers who participated in this study and the local authorities and communities in Equatorial Guinea for their support. We also thank the IPBLN and IBV core facilities for their support to project activities.

## Supporting information

**S1 Text. Tables S1 to S5**

**S6 Table. Genome-based epidemiological analysis of the sequenced samples**

## References

1. World Health Organization. WHO COVID-19 Dashboard [Internet]. [cited 2023 Jul 28]. Available from: https://covid19.who.int/

2. Kannan S, Shaik Syed Ali P, Sheeza A. Omicron (B.1.1.529) - variant of concern-molecular profile and epidemiology: A mini review. Eur Rev Med Pharmacol Sci. 2021;25(24):8019–22.

3. Tegally H, Wilkinson E, Lessells RJ, Giandhari J, Pillay S, Msomi N, et al. Sixteen novel lineages of SARS-CoV-2 in South Africa. Nat Med [Internet]. 2021;27(3):440–6. Available from: 10.1038/s41591-021-01255-3

4. Tegally H, San JE, Cotten M, Moir M, Tegomoh B, Mboowa G, et al. The evolving SARS-CoV-2 epidemic in Africa: Insights from rapidly expanding genomic surveillance. Science [Internet]. 2022 Oct 7 [cited 2023 Jun 5];378(6615). Available from: /pmc/articles/PMC9529057/

5. Hosch S, Mpina M, Nyakurungu E, Borico NS, Obama TMA, Ovona MC, et al. Genomic Surveillance Enables the Identification of Co-infections With Multiple SARS-CoV-2 Lineages in Equatorial Guinea. Front Public Heal. 2022;9(January):1–7.

6. Ozer EA, Simons LM, Adewumi OM, Fowotade AA, Omoruyi EC, Adeniji JA, et al. Multiple expansions of globally uncommon SARS-CoV-2 lineages in Nigeria. Nat Commun. 2022;13(1).

7. MINSABS. Ministerio de Sanidad y Bienestar Social-Guinea Equatorial [Internet]. [cited 2023 Apr 23]. Available from: https://guineasalud.org/estadisticas/

8. Shu Y, McCauley J. GISAID: Global initiative on sharing all influenza data – from vision to reality. Eurosurveillance [Internet]. 2017 Mar 3 [cited 2023 Aug 22];22(13):1. Available from: /pmc/articles/PMC5388101/

9. World Health Organization. Scaling up genomic sequencing in Africa [Internet]. Available from: https://www.afro.who.int/news/scaling-genomic-sequencing-africa

10. Uwishema O, Badri R, Onyeaka H, Okereke M, Akhtar S, Mhanna M, et al. Fighting Tuberculosis in Africa: The Current Situation Amidst the COVID-19 Pandemic. Disaster Med Public Health Prep [Internet]. 2022 [cited 2023 Jul 3]; Available from: https://pubmed.ncbi.nlm.nih.gov/35673793/

11. Shi B, Zheng J, Xia S, Lin S, Wang X, Liu Y, et al. Accessing the syndemic of COVID-19 and malaria intervention in Africa. Infect Dis poverty [Internet]. 2021 Dec 1 [cited 2023 Jul 3];10(1). Available from: https://pubmed.ncbi.nlm.nih.gov/33413680/

12. Uwishema O, Sapkota S, Wellington J, Onyeaka CVP, Onyeaka H. Leishmaniasis control in the light of the COVID-19 pandemic in Africa. Ann Med Surg [Internet]. 2022 Aug 1 [cited 2023 Jul 3];80:104263. Available from: /pmc/articles/PMC9339101/

13. (WHO) WHO. World malaria report 2022. Geneva; 2022.

14. World Health Organization. The potential impact of health service disruptions on the burden of malaria: a modelling analysis for countries in sub-Saharan Africa [Internet]. 2020. 29 p. Available from: https://www.who.int/publications/i/item/9789240004641

15. López-Farfán D, Yerbanga RS, Parres-Mercader M, Torres-Puente M, Gómez-Navarro I, Sanou DMS, et al. Prevalence of SARS-CoV-2 and co-infection with malaria during the first wave of the pandemic (the Burkina Faso case). Front Public Heal [Internet]. 2022 Dec 12 [cited 2023 Jan 18];10. Available from: /pmc/articles/PMC9791192/

16. Amoo OS, Aina OO, Okwuraiwe AP, Onwuamah CK, Shaibu JO, Ige F, et al. COVID-19 Spread Patterns Is Unrelated to Malaria Co-Infections in Lagos, Nigeria. Adv Infect Dis. 2020;10(05):200–15.

17. Muhammad Y, Aminu YK, Ahmad AE, Iliya S, Muhd N, Yahaya M, et al. An elevated 8-isoprostaglandin F2 alpha (8-iso-PGF2α) in COVID-19 subjects co-infected with malaria. Pan Afr Med J [Internet]. 2020 Sep 21 [cited 2022 Sep 15];37(78):1–10. Available from: /pmc/articles/PMC7680236/

18. Onosakponome EO, Wogu MN. The Role of Sex in Malaria-COVID19 Coinfection and Some Associated Factors in Rivers State, Nigeria. J Parasitol Res. 2020;2020.

19. Matangila JR, Nyembu RK, Telo GM, Ngoy CD, Sakobo TM, Massolo JM, et al. Clinical characteristics of COVID-19 patients hospitalized at Clinique Ngaliema, a public hospital in Kinshasa, in the Democratic Republic of Congo: A retrospective cohort study. PLoS One [Internet]. 2020;15(12 December):1–15. Available from: 10.1371/journal.pone.0244272

20. Achan J, Serwanga A, Wanzira H, Kyagulanyi T, Nuwa A, Magumba G, et al. Current malaria infection, previous malaria exposure, and clinical profiles and outcomes of COVID-19 in a setting of high malaria transmission: an exploratory cohort study in Uganda. The Lancet Microbe [Internet]. 2022;3(1):e62–71. Available from: 10.1016/S2666-5247(21)00240-8

21. Bakamutumaho B, Cummings MJ, Owor N, Kayiwa J, Namulondo J, Byaruhanga T, et al. Severe COVID-19 in uganda across two epidemic phases: A prospective cohort study. Am J Trop Med Hyg. 2021;105(3):740–4.

22. Morton B, Barnes KG, Anscombe C, Jere K, Matambo P, Mandolo J, et al. Distinct clinical and immunological profiles of patients with evidence of SARS-CoV-2 infection in sub-Saharan Africa. Nat Commun. 2021;12(1).

23. Hussein R, Guedes M, Ibraheim N, Ali MM, El-Tahir A, Allam N, et al. Impact of COVID-19 and malaria coinfection on clinical outcomes: a retrospective cohort study. Clin Microbiol Infect. 2022 Aug 1;28(8):1152.e1-1152.e6.

24. Ditombi BCM, Ngondza BP, Boulingui CM, Nguema OAM, Ngomo JMN, M’Bondoukwé NP, et al. Malaria and COVID-19 prevalence in a population of febrile children and adolescents living in Libreville. South African J Infect Dis [Internet]. 2022 Oct 26 [cited 2023 Jun 7];37(1). Available from: /pmc/articles/PMC9634652/

25. Sebastião CS, Gaston C, Paixão JP, Euclides |, Sacomboio NM, Neto Z, et al. Coinfection between SARS-CoV-2 and vector-borne diseases in Luanda, Angola. 2021 [cited 2023 Jun 15]; Available from: https://onlinelibrary.wiley.com/doi/10.1002/jmv.27354

26. Ingabire PM, Nantale R, Sserwanja Q, Nakireka S, Musaba MW, Muyinda A, et al. Factors associated with prolonged hospitalization of patients with corona virus disease (COVID-19) in Uganda: a retrospective cohort study. Trop Med Health [Internet]. 2022 Dec 1 [cited 2023 Jun 16];50(1):100. Available from: /pmc/articles/PMC9795158/

27. Mahajan NN, Gajbhiye RK, Bahirat S, Lokhande PD, Mathe A, Rathi S, et al. Co-infection of malaria and early clearance of SARS-CoV-2 in healthcare workers. J Med Virol. 2021;93(4):2431–8.

28. WHO. Malaria Microscopy Quality Assurance Manual. 2009;

29. Grubaugh ND, Gangavarapu K, Quick J, Matteson NL, De Jesus JG, Main BJ, et al. An amplicon-based sequencing framework for accurately measuring intrahost virus diversity using PrimalSeq and iVar. Genome Biol [Internet]. 2019 Jan 8 [cited 2022 Sep 16];20(1):1–19. Available from: https://genomebiology.biomedcentral.com/articles/10.1186/s13059-018-1618-7

30. Wood DE, Salzberg SL. Kraken: Ultrafast metagenomic sequence classification using exact alignments. Genome Biol [Internet]. 2014 Mar 3 [cited 2022 Sep 16];15(3):1–12. Available from: https://genomebiology.biomedcentral.com/articles/10.1186/gb-2014-15-3-r46

31. Chen S, Zhou Y, Chen Y, Gu J. fastp: an ultra-fast all-in-one FASTQ preprocessor. Bioinformatics [Internet]. 2018 Sep 9 [cited 2022 Sep 16];34(17):i884. Available from: /pmc/articles/PMC6129281/

32. Ewels P, Magnusson M, Lundin S, Käller M. MultiQC: summarize analysis results for multiple tools and samples in a single report. Bioinformatics [Internet]. 2016 Oct 1 [cited 2022 Sep 16];32(19):3047–8. Available from: https://academic.oup.com/bioinformatics/article/32/19/3047/2196507

33. Wu F, Zhao S, Yu B, Chen YM, Wang W, Song ZG, et al. A new coronavirus associated with human respiratory disease in China. Nat 2020 5797798 [Internet]. 2020 Feb 3 [cited 2022 Sep 16];579(7798):265–9. Available from: https://www.nature.com/articles/s41586-020-2008-3

34. Katoh K, Standley DM. MAFFT Multiple Sequence Alignment Software Version 7: Improvements in Performance and Usability. Mol Biol Evol [Internet]. 2013 Apr 1 [cited 2022 Sep 16];30(4):772–80. Available from: https://academic.oup.com/mbe/article/30/4/772/1073398

35. O’Toole Á, Scher E, Underwood A, Jackson B, Hill V, McCrone JT, et al. Assignment of epidemiological lineages in an emerging pandemic using the pangolin tool. Virus Evol [Internet]. 2021 [cited 2022 Sep 16];7(2). Available from: /pmc/articles/PMC8344591/

36. Aksamentov I, Roemer C, Hodcroft EB, Neher RA. Nextclade: clade assignment, mutation calling and quality control for viral genomes. J Open Source Softw [Internet]. 2021 Nov 30 [cited 2022 Sep 16];6(67):3773. Available from: https://joss.theoj.org/papers/10.21105/joss.03773

37. Hadfield J, Megill C, Bell SM, Huddleston J, Potter B, Callender C, et al. NextStrain: Real-time tracking of pathogen evolution. Bioinformatics. 2018 Dec 1;34(23):4121–3.

38. Sagulenko P, Puller V, Neher RA. TreeTime: Maximum-likelihood phylodynamic analysis. Virus Evol [Internet]. 2018 Jan 1 [cited 2022 Sep 16];4(1). Available from: /pmc/articles/PMC5758920/

39. Martínez-Martínez FJ, Massinga AJ, De Jesus Á, Ernesto RM, Cano-Jiménez P, Chiner-Oms Á, et al. Tracking SARS-CoV-2 introductions in Mozambique using pandemic-scale phylogenies: a retrospective observational study. Lancet Glob Heal. 2023;11(6):e933–41.

40. Turakhia Y, Thornlow B, Hinrichs AS, Maio N, Gozashti L, Lanfear R, et al. Ultrafast Sample placement on Existing tRees (UShER) enables real-time phylogenetics for the SARS-CoV-2 pandemic. Nat Genet [Internet]. [cited 2023 Aug 23]; Available from: 10.1038/s41588-021-00862-7

41. Lanfear R. Global phylogenies of SARS-CoV-2 sequences [Internet]. Available from: https://github.com/roblanf/sarscov2phylo

42. Thornlow B. matUtils [Internet]. Available from: https://usher-wiki.readthedocs.io/en/latest/matUtils.html

43. Sanderson T. Taxonium, a web-based tool for exploring large phylogenetic trees eLife 11:e82392. Elife [Internet]. 2022;11:e82392. Available from: 605 10.7554/eLife.82392%0A

44. Kusi KA, Frimpong A, Partey FD, Lamptey H, Amoah LE, Ofori MF. High infectious disease burden as a basis for the observed high frequency of asymptomatic SARS-CoV-2 infections in sub-Saharan Africa. AAS Open Res. 2021;4(May):2.

45. Hussein MIH, Albashir AAD, Elawad OAMA, Homeida A. Malaria and COVID-19: unmasking their ties. Malar J [Internet]. 2020;19(1):1–10. Available from: 10.1186/s12936-020-03541-w

46. Osei SA, Biney RP, Anning AS, Nortey LN, Ghartey-Kwansah G. Low incidence of COVID-19 case severity and mortality in Africa; Could malaria co-infection provide the missing link? BMC Infect Dis [Internet]. 2022;22(1):1–11. Available from: 10.1186/s12879-022-07064-4

47. Iesa MAM, Osman MEM, Hassan MA, Dirar AIA, Abuzeid N, Mancuso JJ, et al. SARS-CoV-2 and Plasmodium falciparum common immunodominant regions may explain low COVID-19 incidence in the malaria-endemic belt. New microbes new Infect [Internet]. 2020 Nov 1 [cited 2022 Jul 7];38. Available from: https://pubmed.ncbi.nlm.nih.gov/33230417/

48. Raham TF. Influence of malaria endemicity and tuberculosis prevalence on COVID-19 mortality. Public Health [Internet]. 2021;194:33–5. Available from: 10.1016/j.puhe.2021.02.018

49. Johns Hopkins University. Johns Hopkins coronavirus resource center [Internet]. [cited 2023 Jul 10]. Available from: https://coronavirus.jhu.edu/region/equatorial-guinea

50. Wilkinson E, Giovanetti M, Tegally H, San JE, Lessells R, Cuadros D, et al. A year of genomic surveillance reveals how the SARS-CoV-2 pandemic unfolded in Africa. Science (80-). 2021;374(6566):423–31.

51. cov-lineages.org [Internet]. Available from: https://cov-lineages.org/lineage_list.html

52. Mlcochova P, Kemp SA, Dhar MS, Papa G, Meng B, Ferreira IATM, et al. SARS-CoV-2 B.1.617.2 Delta variant replication and immune evasion. Nat 2021 5997883 [Internet]. 2021 Sep 6 [cited 2023 Jun 5];599(7883):114–9. Available from: https://www.nature.com/articles/s41586-021-03944-y

53. Martínez-Martínez J, Cano-Jiménez P, Chiner-Oms Á, Bsc G-N, Guillot-Fernández M, Jiménez-Serrano S, et al. Tracking SARS-CoV-2 introductions in Mozambique using pandemic-scale phylogenies: a retrospective observational study. Artic Lancet Glob Heal [Internet]. 2023 [cited 2023 Aug 23];11:933–74. Available from: https://gitlab.

54. López-Farfán D, Irigoyen N, Gómez-Díaz E. Exploring SARS-CoV-2 and Plasmodium falciparum coinfection in human erythrocytes. Front Immunol [Internet]. 2023 Mar 13 [cited 2023 Jun 5];14. Available from: https://pubmed.ncbi.nlm.nih.gov/36993979/

55. Huerga Encabo H, Grey W, Garcia-Albornoz M, Wood H, Ulferts R, Aramburu IV, et al. Human Erythroid Progenitors Are Directly Infected by SARS-CoV-2: Implications for Emerging Erythropoiesis in Severe COVID-19 Patients. Stem cell reports [Internet]. 2021 Mar 9 [cited 2022 Aug 10];16(3):428–36. Available from: https://pubmed.ncbi.nlm.nih.gov/33581053/

56. Shahbaz S, Xu L, Osman M, Sligl W, Shields J, Joyce M, et al. Erythroid precursors and progenitors suppress adaptive immunity and get invaded by SARS-CoV-2. Stem Cell Reports. 2021 May 11;16(5):1165–81.

